# Altered prefrontal signaling during inhibitory control in a salient drug context in human cocaine addiction

**DOI:** 10.1101/2021.09.27.21264113

**Authors:** Ahmet O. Ceceli, Muhammad A. Parvaz, Sarah King, Matthew Schafer, Pias Malaker, Akarsh Sharma, Nelly Alia-Klein, Rita Z. Goldstein

## Abstract

Drug addiction is characterized by impaired Response Inhibition and Salience Attribution (iRISA), where the salience of drug cues is postulated to overpower that of other reinforcers with a concomitant decrease in self-control. However, the neural underpinnings of the interaction between the salience of drug cues and inhibitory control in drug addiction remain unclear. We developed a novel stop-signal fMRI task where the stop-signal reaction time (SSRT—a classical inhibitory control measure) was tested under different salience conditions (modulated by drug, food, threat or neutral words) in individuals with cocaine use disorder (CUD; n=26) vs. demographically matched healthy control participants (HC; n=26). Despite similarities in drug cue-related SSRT and valence and arousal word ratings between groups, dorsolateral prefrontal cortex (dlPFC) activity was diminished during the successful inhibition of drug versus food cues in CUD, and was correlated with lower frequency of recent use, lower craving, and longer abstinence (Z>3.1, *p*<.05 corrected). Results suggest altered involvement of cognitive control regions (e.g., dlPFC) during inhibitory control under a drug context, relative to an alternative reinforcer, in CUD. Supporting the iRISA model, these results elucidate the direct impact of drug-related cue-reactivity on the neural signature of inhibitory control in drug addiction.

---

The impaired response inhibition and salience attribution (iRISA) model of drug addiction posits that addicted individuals attribute excessive salience to drug and drug-related cues at the expense of non-drug-related reinforcers, with concomitant decreases in inhibitory control (Goldstein and Volkow 2002, 2011). Consistent with the animal literature (Phillips et al. 2003), in human neuroimaging studies increased responses in the mesencephalon (Goldstein, Tomasi, et al. 2009), anterior cingulate cortex and ventromedial prefrontal cortex (vmPFC; Kober et al., 2016; Konova et al., 2019), and connectivity between salience, reward, and executive networks (Ray et al. 2015), have been interpreted to reflect the excessive salience attributed to drug cues in drug-addicted individuals. Inhibitory control (in non-drug contexts) has instead been associated with hypoactivations in the inferior frontal gyrus (IFG), orbitofrontal cortex (OFC), and dorsolateral PFC (dlPFC) in this population as previously reviewed (Zilverstand et al. 2018; Ceceli et al. 2021). However, the interaction between inhibitory control and drug cue reactivity, crucial for more accurately examining the neurobehavioral bases of drug addiction’s core phenomenology, has yet to be well-characterized.

Several neuroimaging studies in addicted individuals have used drug related vs. non-drug related cues in the context of tasks approximating inhibitory control such as the Go/No-Go (Ames et al. 2014; Czapla et al. 2017; Gilman et al. 2018) and Stroop tasks (Carpenter et al. 2006; Hester et al. 2006; Goldstein et al. 2007; Goldstein, Alia-Klein, et al. 2009; Goldstein, Tomasi, et al. 2009; Smith and Ersche 2014; DeVito et al. 2018). Using the Go/No-Go task, increased activations in the dlPFC, vmPFC, and insula during alcohol-related No-Go vs. non-alcohol Go trials have been reported in alcohol dependent subjects (Ames et al. 2014; Czapla et al. 2017) [with negative results in smokers (Gilman et al. 2018)]. Instead of measuring the classical Stroop conflict induced by reading (faster) vs. naming (slower) of incongruent color words (Jensen and Rohwer 1966), the drug-Stroop task measures the attentional interference induced by color-naming of drug vs. non-drug related words. This drug-related attentional interference manifests as prolonged response times (Ersche et al. 2010) and/or altered neural activation in the brain’s reward/salience circuitry [OFC, anterior cingulate, striatum, and midbrain (Goldstein, Alia-Klein, et al. 2009; Goldstein, Tomasi, et al. 2009; Smith and Ersche 2014)]. However, while the Go/No-Go task captures response selection (Raud et al. 2020), it does not measure the ability to stop after response initiation, and the drug-Stroop task does not create an inherent inhibitory control demand, rendering both task types insufficient in informing potential lapses in self-control under a motivationally challenging state, such as craving.

The stop-signal task (SST) permits the investigation of inhibitory control by estimating the competition between the “Go” and “Stop” processes (Verbruggen and Logan 2008), and has previously been used to demonstrate the neural correlates of inhibitory control in drug addiction (Li et al. 2008, 2009; Matuskey et al. 2013; Elton et al. 2014; Sjoerds et al. 2014; Hu et al. 2015; Harlé et al. 2016, 2019; Wang et al. 2018; Zhang et al. 2018; Sakoglu et al. 2019; Zhukovsky et al. 2021). However, a gap remains in testing the potential modulation of inhibitory control by drug cue salience for the inspection of their interaction at the brain level. In individuals with cocaine use disorder (CUD) and matched healthy control (HC) subjects, we present results of a novel SST that allowed for the parametric/trial-by-trial modulation of inhibitory control by salient drug, other salient non-drug (e.g., food), and neutral cues (words) during fMRI. Further, we conservatively pre-selected our data to abide by recent recommendations of best practices in estimating stopping ability (Verbruggen et al. 2019). We probed three core hypotheses: 1) Regardless of cue type and group, prefrontal regions would drive inhibitory control processes on this hybrid task; 2) inhibitory control-related prefrontal signaling would be altered in the CUD group; and 3) the dlPFC (and other inhibitory PFC regions, e.g., the IFG) would be crucial for inhibitory control under drug cue reactivity in CUD, and this effect would be supported by a link with cocaine use severity.

## Materials and Methods

### Participants

Participants were recruited using flyers, newspaper ads, and by word of mouth. From a dataset of 79 participants with valid behavioral data (i.e., behavioral task performance that permitted an estimation of inhibitory control via SSRT), we pre-selected 52 age-, education-, and verbal IQ-matched [using the R package *MatchIt* (Ho et al. 2011)], right-handed (given the verbal nature of this task and concerns about lateralization differences, as estimated with the Edinburgh Handedness Inventory score > 75%) individuals with complete behavioral and fMRI data (n=26 per group, 43 men, mean age: 43.4 ± 7.6; see *Behavioral data analysis* and *BOLD-fMRI data analysis* sections for details, and Table 1 for sample profile).

**Table 1.** Sample profile.

|  | CUD (n = 26) | HC (n = 26) | Significance test |
| --- | --- | --- | --- |
| Age | 44.07 ± 8.18<br>Min: 28, Max: 59 | 42.66 ± 7.05<br>Min: 27, Max: 55 | $t(50) = 0.67, p = .509$ |
| Sex (M/F) | 22/4 | 21/5 | $\chi^2(1) = 0.13, p = .718$ |
| Race (Black/White/Other/Unreported) | 17/3/4/2 | 16/4/6/0 | $\chi^2(2) = 1.69, p = .430$ |
| Education (years; 12=high school grad) | 12.27 ± 1.59<br>Min: 9, Max: 16 | 12.92 ± 1.72<br>Min: 10, Max: 16 | $t(50) = 1.42, p = .160$ |
| Verbal IQ (std. WRAT-III score) | 95.04 ± 11.65<br>Min: 76, Max: 116 | 99.00 ± 11.30<br>Min: 75, Max: 113 | $t(50) = 1.24, p = .219$ |
| Non-verbal IQ (matrix reasoning score) | 8.96 ± 2.99<br>Min: 3, Max: 14 | 11.12 ± 2.16<br>Min: 4, Max: 15 | $t(50) = 2.98, p = .004^*$ |
| Cigarette use per day (cigarettes) | 5.11 ± 6.20<br>Min: 0, Max: 25 | 2.92 ± 6.21<br>Min: 0, Max: 20 | $t(50) = 1.27, p = .209$ |
| Depression (BDI score) | 7.81 ± 7.65<br>Min: 0, Max: 24 | 4.46 ± 5.69<br>Min: 0, Max: 20 | $t(50) = 1.79, p = .079$ |
| Duration of cocaine use (years) | 16.00 ± 8.15<br>Min: 3, Max: 30 | -- | -- |
| Past month cocaine use (days/month) | 6.08 ± 8.34<br>Min: 0, Max: 25 | -- | -- |
| Days since last cocaine use | 293.23 ± 530.65<br>Min: 0, Max: 1825 | -- | -- |
| Cocaine Severity of Dependence Scale | 5.04 ± 4.87<br>Min: 0, Max: 15 | -- | -- |
| Cocaine Selective Severity Assessment | 16.35 ± 11.33<br>Min: 0, Max: 37 | -- | -- |
| Cocaine Craving Questionnaire | 13.73 ± 13.78<br>Min: 0, Max: 45 | -- | -- |

Note: Significant group differences are flagged with an asterisk. ± denotes standard deviation.

Participants underwent a series of clinical and neuropsychological assessments delivered by trained staff under a clinical psychologist’s supervision. These interviews included the Structured Clinical Interview for DSM IV for Axis I Disorders (American Psychiatric Association 2013); the Addiction Severity Index (McLellan et al. 1992)—a semi-structured interview that assesses the severity as well as recent and lifetime history of alcohol- and drug-related problems; the Cocaine Selective Severity Assessment (Kampman et al. 1998) for evaluating cocaine abstinence/withdrawal signs and symptoms (i.e., sleep impairment, anxiety, energy levels, craving and depressive symptoms) 24 hours within time of interview; the 5-item Cocaine Craving Questionnaire (Tiffany et al. 1993); and the 5-item Severity of Dependence Scale (Gossop et al. 1992). A brief physical examination encompassing height, weight, urine drug toxicology, breath alcohol and carbon monoxide (for recent cigarette use) levels, and medical history, were also obtained by trained research staff.

Participants were excluded for the following: 1) history of major psychiatric disorders [barring other substance use disorders and highly comorbid illnesses such as post-traumatic stress disorder (PTSD) for CUD, and nicotine dependence for HC]; 2) history of seizures or other central nervous system disorders; 3) cardiovascular, endocrinological, metabolic, oncological, or autoimmune disease; 4) MRI contraindications; 5) presence of any psychoactive drugs or their metabolites (except for cocaine in the CUD group) in urine assays; 6) evidence of alcohol and/or any other drug-related intoxication at the time of participation; 7) positive pregnancy test (determined via a urine assay); and 8) color blindness [assessed with the Ishihara color blindness test (Clark 1924)].

All participants in the CUD group met criteria for substance use disorder (with cocaine and/or crack cocaine as the primary substance of choice, 23 meeting criteria for dependence, and three for abuse). Urine screen results confirmed the presence of cocaine in 22 participants in the CUD group. Urine screens for all other drugs were negative (the HC participants tested negative for all drugs, including cocaine). Comorbid diagnoses within CUD included current intermittent explosive disorder (N = 5), PTSD in full remission (N = 1), specific phobias (N = 2), marijuana dependence (N = 13), alcohol abuse and/or dependence (N = 26), opioid dependence (N = 8), amphetamine dependence (N = 1), hallucinogen abuse and/or dependence (N = 2), and polysubstance dependence (N = 3). Diagnoses among HC included intermittent explosive disorder (current: N = 10, past: N = 1), PTSD in full remission (N = 2), specific phobias (N = 1), marijuana abuse and/or dependence (N = 3) and alcohol dependence (N = 6). All substance use comorbidities were in full remission (except for one CUD participant who met criteria for marijuana dependence, partial remission). Participants received full information about the research and provided written consent in accordance with the institutional review board of the Icahn School of Medicine at Mount Sinai and Code of Ethics of the World Medical Association [Declaration of Helsinki (Rickham 1964)].

### Experimental Design and Statistical Analysis

#### Emotional Stop-Signal Task

In the traditional SST, participants are instructed to respond to a frequent neutral Go signal as quickly as possible, and inhibit responses to this cue when it is followed by a rare stop-signal [e.g., a visual cue or an auditory tone (Verbruggen and Logan 2008) superimposed on the Go signal; see Figure 1, left].

**Figure 1.**
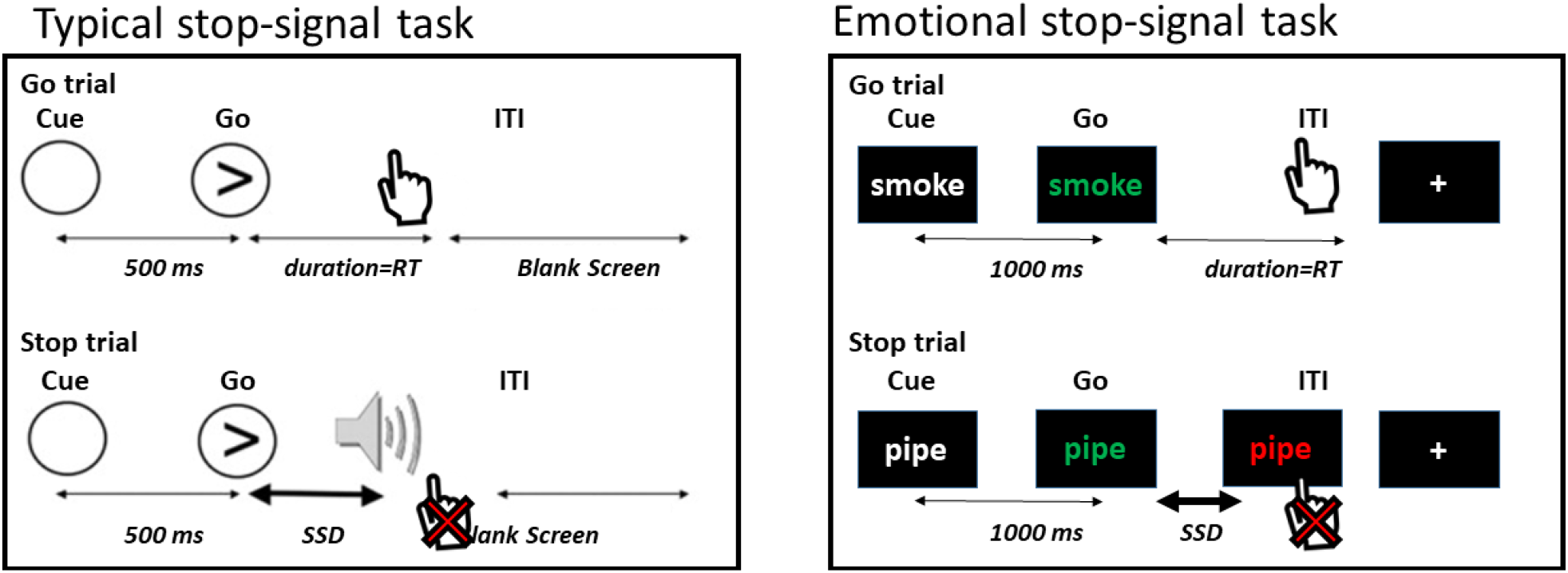
The Emotional Stop-Signal Task. Participants are instructed to respond via button presses as quickly and accurately as possible to the change in word color from white to blue/green, and to suppress their responses when the word color turns red after a variable delay (i.e., the stop-signal delay, or SSD). Uniquely, word categories comprised neutral words, as well as drug, food, and threat-related words, permitting the examination of emotional reactivity during inhibitory control. Typical SST figure adapted from Aron and Poldrack, 2006.

Here, during fMRI, we presented drug, food, and threat-related words in addition to neutral words as the Go signals in a modified emotional SST (see Figure 1, right). These words were matched in length and frequency in the English language [mean frequency: drug words = 23.69, food words = 15.70, threat words = 19.87, neutral words = 21.97; (Francis et al. 1982)]. Each trial started with a fixation cross that jittered in duration between 2000 and 2600 msec, which was then followed by a drug, food, threat, or neutral word in white text over a black background (1000 msec). Next, the word in white text changed in color to blue or green, to which the participants were instructed to respond with their right (for blue font) or left (for green font) index finger using MR compatible response gloves. These Go trials comprised 50% of trials; in 25% of trials, a stop-signal followed the change in color, such that the blue or green font turned red after a variable delay (i.e., the stop-signal delay, or SSD). This change instructed the participants to suppress their response (when the word color turned red; see Figure 1, right bottom). The SSD was set to an initial duration of 250 msec and adjusted in parallel to the participant’s stopping ability for each cue type (yielding four independent SSD adjustments to precisely estimate stopping speed to each cue). When the participant successfully stopped, the SSD increased by 50 msec (making the next stop trial more difficult), and when the participant failed to stop, the SSD decreased by 50 msec (making the next stop trial easier). The remaining 25% of trials were empty trials in the form of a fixation cross (2 sec; same as the Go trial duration) included to minimize anticipatory effects and improve signal detection (Hagberg et al. 2001; Wager and Nichols 2003). Participants underwent four runs of this task, each containing 48 Go and 24 Stop trials (18 trials in each word category per run) for a total of 192 Go trials, 96 Stop trials (and 72 trials in each word category). After the MRI session, participants were instructed to rate each word using a 5-point scale on valence [“Please rate how you currently feel about the above words from PLEASANT (leftmost) to UNPLEASANT (rightmost) using the scale.”] and arousal [“Please rate how emotional you currently feel about the above words from VERY EMOTIONAL (leftmost) to CALM (rightmost) using the scale.”].

#### MRI data acquisition

Scanning was conducted on a Siemens 3T Skyra (Siemens, Erlangen, Germany), using a 32-channel head coil. The BOLD-fMRI responses were measured as a function of time using a T2*-weighted single shot multi-band accelerated gradient-echo EPI sequence [TE/TR=35/1000 msec, 2.1 mm isotropic resolution, no gap, 70 axial slices for whole brain (14.7 cm) coverage, FOV 206 × 181 mm, matrix size 96 × 84, 60°-flip angle (approximately Ernst angle), multi-band factor of 7, blipped CAIPIRINHA phase-encoding shift=FOV/3, ∼2 kHz/Pixel bandwidth with ramp sampling, echo spacing 0.68 msec, and echo train length 57.1 msec]. Each of the four fMRI task runs were approximately 6 min 30 sec in length, totaling about 26 min. T1-weighted anatomical images were acquired using a 3D MPRAGE sequence [FOV 256 × 256 × 179 mm, 0.8 mm^3^ isotropic resolution, TR/TE/TI=2400/2.07/1000 msec, flip angle 8° with binomial (1, −1) fat saturation, bandwidth 240 Hz/pixel, echo spacing 7.6 msec, and in-plane acceleration (GRAPPA) factor of 2, with a total acquisition time of ∼7 min]. The 90-min scan session included additional procedures (presented in a randomized order to circumvent possible order effects) reported elsewhere (Moeller et al. 2018).

#### MRI data preprocessing

Raw fMRI data in DICOM format were converted to NIFTI using dcm2niix (Li et al. 2016) and adapted to Brain Imaging Data Structure (BIDS) standards to enhance neuroimaging data portability and reproducibility (Gorgolewski et al. 2016). BIDS-validated data were then preprocessed via the Nipype-based fMRIPrep pipeline (version 1.5.0) (Gorgolewski et al. 2011; Esteban et al. 2019). FMRIPrep is a robust fMRI preprocessing pipeline that recruits tools from well-established neuroimaging software (e.g., FSL, Freesurfer, AFNI) to standardize and optimize fMRI preprocessing (Esteban et al. 2019). FMRIPrep’s workflow is summarized as follows: structural images were corrected for intensity non-uniformity and skull-stripped using ANTS (Tustison et al. 2010). These structural images were spatially normalized to the ICBM 152 Nonlinear Asymmetrical template via nonlinear registration using ANTS (Avants et al. 2008; Fonov et al. 2009). Brain tissue was segmented into white matter, gray matter, and cerebrospinal fluid using FSL’s FAST (Zhang et al. 2001). Functional data were corrected for motion artifacts using FSL’s MCFLIRT and for distortion using spin-echo field maps acquired in opposing phase encoding directions via AFNI’s 3dQwarp (Cox 1996; Jenkinson et al. 2002). Motion and distortion corrected images were then co-registered to the participant’s structural images using boundary-based registration with 9 degrees of freedom via FSL’s FLIRT (Jenkinson and Smith 2001; Fonov et al. 2009). These correction, transformation, and registration steps were integrated into a single step transformation workflow using ANTS. In addition to the fMRIPrep workflow, we identified volumes with spikes in translation and rotation parameters in relation to a reference volume. Using a typical boxplot threshold (75th percentile + 1.5 * interquartile range) via FSL’s *fsl_motion_outliers* package, we regressed out an average of 5.92% of outlier volumes in each run (range: 0.51%-14.32%). Average number of outlier volumes regressed out from participants in the CUD group did not significantly differ from those in the HC group, *t*(50) = 0.71, *p* = .483. As a result, there remained no substantial volume-to-volume movement (framewise displacement was below voxel dimensions at mean motion: 0.05 mm; maximum motion: 0.17 mm), with no significant differences across groups, *t*(50) = 0.61, *p* = .542. The preprocessed data from the fMRIPrep pipeline were filtered with a high-pass filter (100 s cutoff) to ignore scanner drift, and spatially smoothed using a Gaussian kernel (5 mm full-width at half maximum) to improve signal to noise ratio. We set an a priori threshold for mean voxel intensity values in all preprocessed images for quality control: BOLD runs with mean voxel intensity at 3 standard deviations above or below the mean of the sample were removed from analyses to mitigate the potential effects of scanner artifacts (excluded CUD n = 2).

#### Behavioral data analysis

A core assumption of the stop-signal paradigm is that Go and Stop processes are in competition to exert control over motor output—a phenomenon described by the horse-race model of response inhibition (Logan and Cowan 1984). To ensure that we accurately estimated stopping ability using our novel task, we followed a recently documented consensus guide for upholding the validity of the horse-race model while analyzing SST data (Verbruggen et al. 2019). We used the integration method to calculate SSRT, in that we identified the nth RT in the Go RT distribution, with n being the number of Go RTs multiplied by the proportion of trials where a response was made to a stop-signal. The nth represents the finishing time of the stopping process (and improves on the common “mean RT” method) (Verbruggen et al. 2019). Accordingly, we only included data from task runs in which the participant displayed mean Go accuracy of 60% or higher, mean Stop accuracy greater than or equal to 25% but less than or equal to 75% (ensuring that the SSD indeed resulted in a competition between Go and Stop processes), and positive mean SSRT. After removing task runs in which the above criteria were violated, there were no significant differences between groups in number of valid task runs [*t*(77) = 0.45, *p* = .652].

We performed a mixed-design repeated measures ANOVA, with SSRT as the dependent variable, Group (CUD, HC) as the between- and Cue (Drug, Food, Threat, Neutral) as the within-subject factors. We followed up on significant interactions with post-hoc t-tests where appropriate. Because the data were pre-selected based on Go and Stop accuracy performance (needed by definition to ascertain validity of the use of this task as an inhibitory control measure), we did not focus on statistical differences based on these variables. To inspect for potential covariates and the impact of clinical measures, select demographics and drug use severity measures were used in Spearmen’s correlational analyses with average and Drug SSRT, Holm-corrected for multiple comparisons. Post-task ratings to the word cues were entered into two-way ANOVAs (Group as between-, Cue as within-subject factors, valence and arousal as separate dependent variables) to test whether participants differentially perceived the valence and arousal of the words across cue types and groups.

#### BOLD-*fMRI data analysis*

Parameter estimates were generated for each participant using the general linear model (GLM) approach via FSL’s FEAT (version 5.98; Woolrich et al., 2001). Four events (Go_Success for successful Go responses, Go_Fail for missed Go trials or response selection errors, Stop_Success for successfully inhibited responses following a stop-signal, and Stop_Fail for failed inhibitions following a stop-signal) per cue type (drug, food, threat, and neutral) were modeled, yielding 16 cue-specific regressors (e.g., Drug_Stop_Success, Food_Stop_Fail, etc.). Regressors were sampled from the onset of the corresponding trials’ Go signals using 1 sec events and convolved with a canonical hemodynamic response function along with their temporal derivatives to be entered into the GLM. Fixation events were not modeled, in that they contributed to the task baseline for linear contrasts—a practice that is in line with prior analyses of the SST (Aron and Poldrack 2006). We used each cue’s Go_Fail events and the *fsl_motion_outlier* outputs denoting volumes with excessive spikes in intensity as regressors of no interest. We removed individuals who did not yield complete fMRI regressors to represent behavioral performance in line with the horse-race model assumptions. For example, if a participant’s non-missing task runs were insufficient in yielding all the necessary task events (e.g., a missing Drug_Stop_Success because the participant had no successful stops during Drug cues in their only remaining run), we excluded the participant from analyses. We based our behavioral and neural analyses on a final pool of 52 participants for consistency and examinations of behavioral performance vis-à-vis BOLD activity (see Table 1 for sample profile).

In each of the four runs, the first level analyses comprised all regressors’ contrasts with baseline (e.g., Drug_Go_Success > Baseline to examine Go-related processes in drug cue trials). Importantly, we used Stop_Success > Stop_Fail as the hallmark contrast of inhibitory control. Thus, the first-level GLM included contrasts for detecting cue-general (Stop_Success > Stop_Fail regardless of cue type) and cue-specific (e.g., Drug_Stop_Success > Drug_Stop_Fail) inhibitory control processes. We contrasted drug with food—a competing, non-drug positively valenced reinforcer [i.e., (Drug_Stop_Success > Drug_Stop_Fail) > (Food_Stop_Success > Food_Stop_Fail)] to focus specifically on regulation of salient drug versus non-drug-related inhibitory control—a practice with precedence and specificity to predicting drug use in CUD (Martinez et al. 2009; Moeller et al. 2009, 2010, 2018). We also contrasted drug-related inhibition with the non-salient neutral category of cues [i.e., (Drug_Stop_Success > Drug_Stop_Fail) > (Neutral_Stop_Success > Neutral_Stop_Fail)], permitting the examination of drug-related arousal. These contrasts in each run were entered into a fixed-effects model to generate aggregate parameter estimates representing general and cue-specific inhibitory control processes at the participant level.

Other iterations of cue contrasts (e.g., Food > Threat, Drug > Threat) were also modeled for consistency, although we did not have an a priori interest in these comparisons for the current purposes. In other words, although we used in the GLM threat cues as the non-drug category that accounts for the potentially negative valence of drug cues, direct comparisons involving threat are outside the scope of our primary interests regarding the iRISA interaction for the current purposes.

To investigate the neural mechanisms of inhibitory control across the CUD and HC groups, the resulting participant-level activation maps were analyzed in a higher level analysis using FSL’s FLAME 1 & 2 (FMRIB’s Local Analysis of Mixed Effects), which improves group-level variance estimates via Markov Chain Monte Carlo simulations to allow for population inferences (Beckmann et al. 2003). We chose a cluster defining threshold of *p* < .001, corrected to a cluster-extent threshold of *p* < .05, in line with common practices to minimize Type I error (Eklund et al. 2016).

To detect cue-general inhibitory control related neural activation associated with overall stopping ability in the task, we performed a similar higher-level analysis with identical thresholding and variance estimation methods, with the addition of mean SSRT as a covariate. We also included cue-specific SSRT covariates that mirrored the BOLD contrasts for cue comparisons. For instance, a covariate containing “Drug SSRT minus Food SSRT” values was created to detect activation patterns during drug versus food inhibition that varied based on behavioral performance to drug versus food cues. Finally, we used select drug use severity measures (i.e., years of cocaine use, frequency of use in past 30 days, days since last use, severity of dependence, withdrawal, and craving) to reveal whether these estimates correlated with any regional activation in the brain during drug cue-related inhibitory control.

## Results

### Behavioral results

The mixed-design repeated measures ANOVA of SSRT revealed no main effect of Group, *F*(1,50) = 3.13, *p* = .083, no main effect of Cue, *F*(3,150) = 0.65, *p* = .584, but a significant Group x Cue interaction, *F*(3,150) = 3.15, *p* = .027, where HC exhibited prolonged SSRT to Food and Neutral cues (see Figure 2 and Table 2). Descriptive statistics for Go and Stop accuracy and Go RT-related group differences are included in Table 2.

**Table 2.**
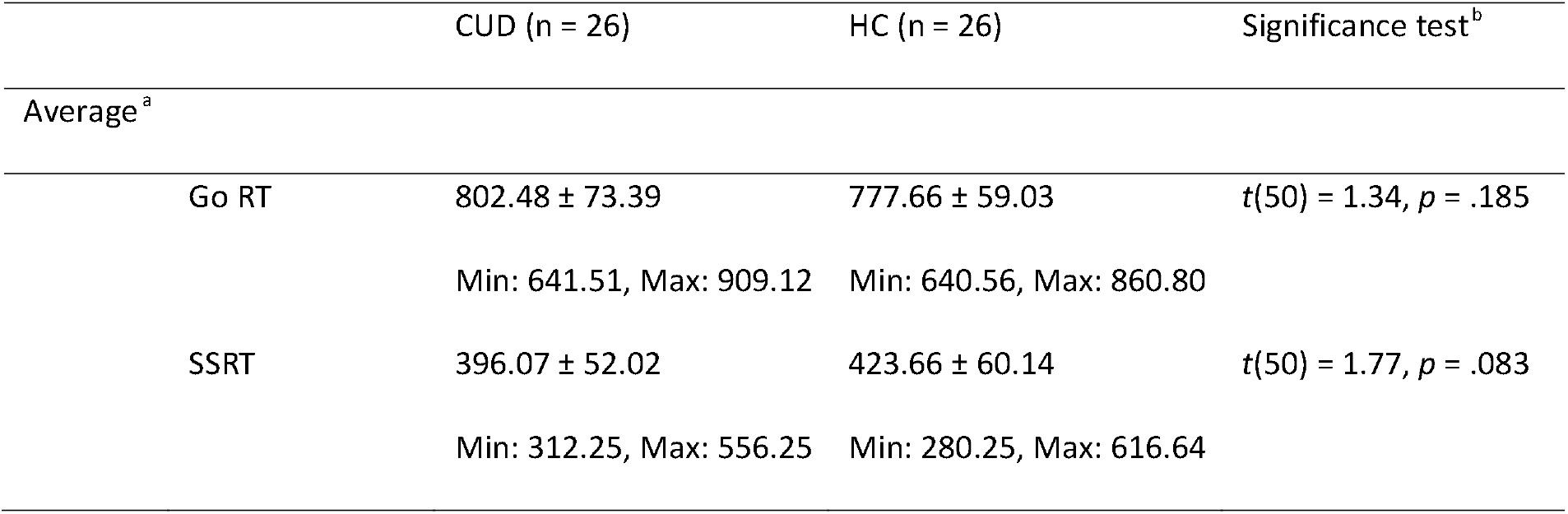
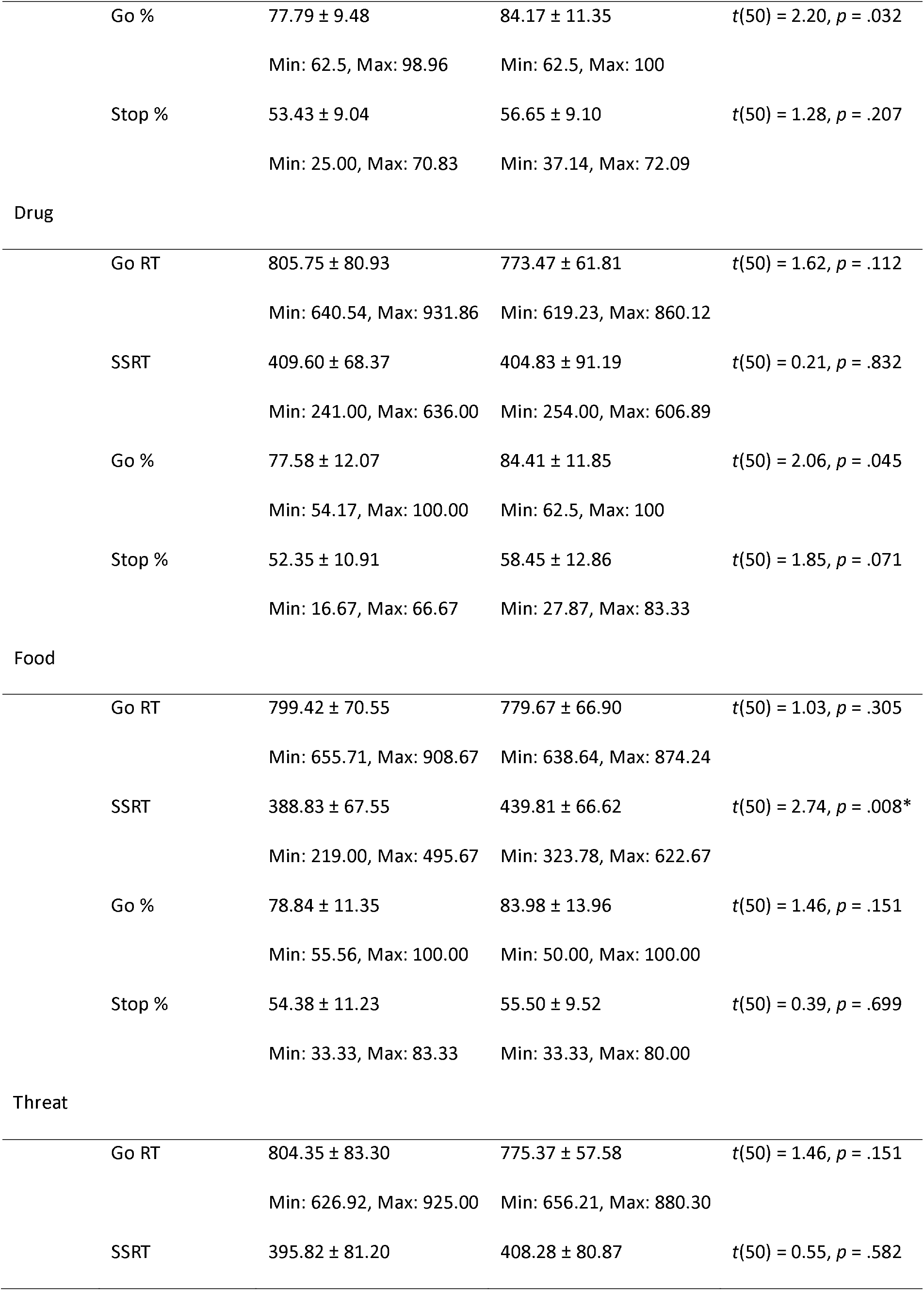
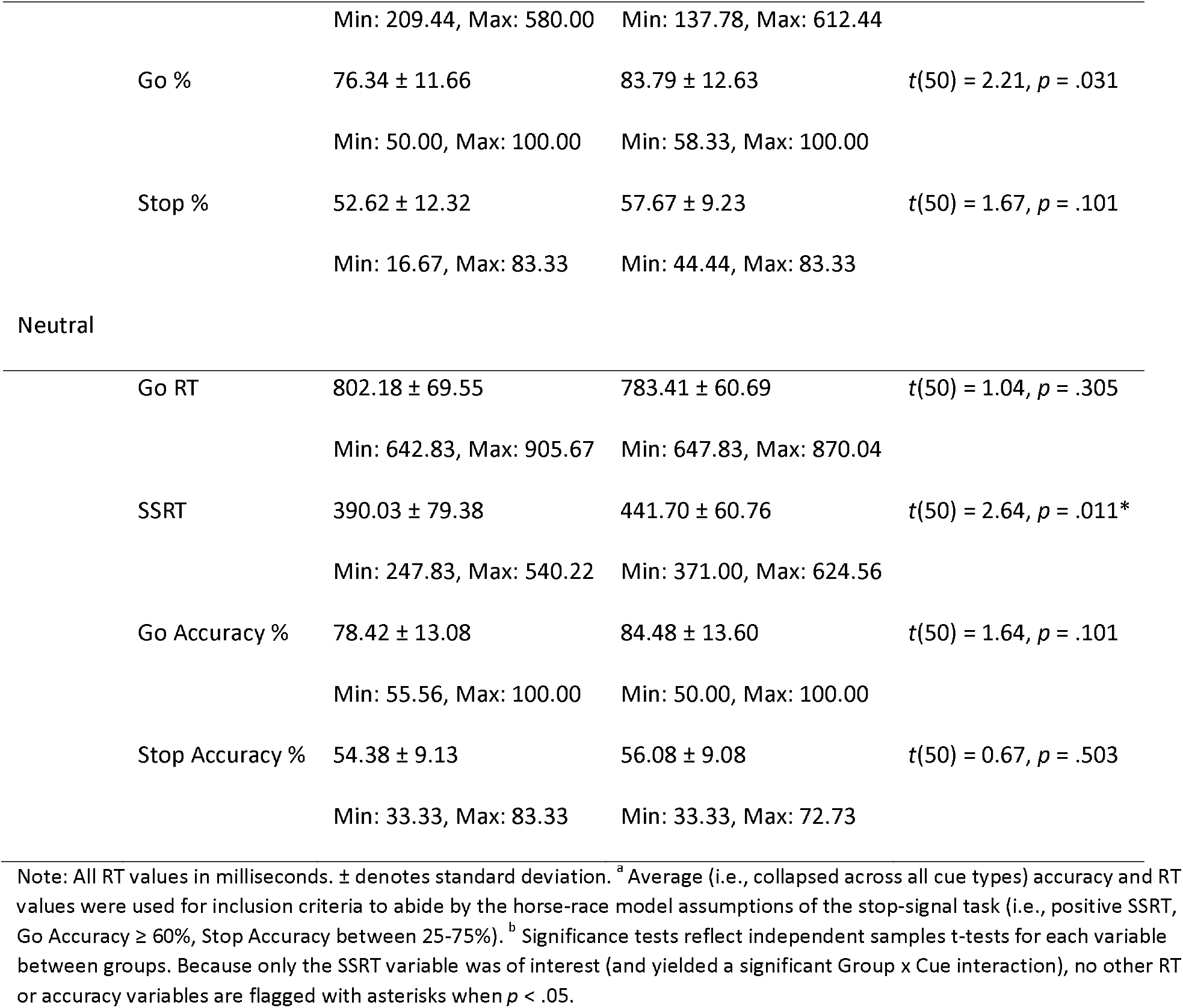
Behavioral inhibitory control performance.

**Figure 2.**
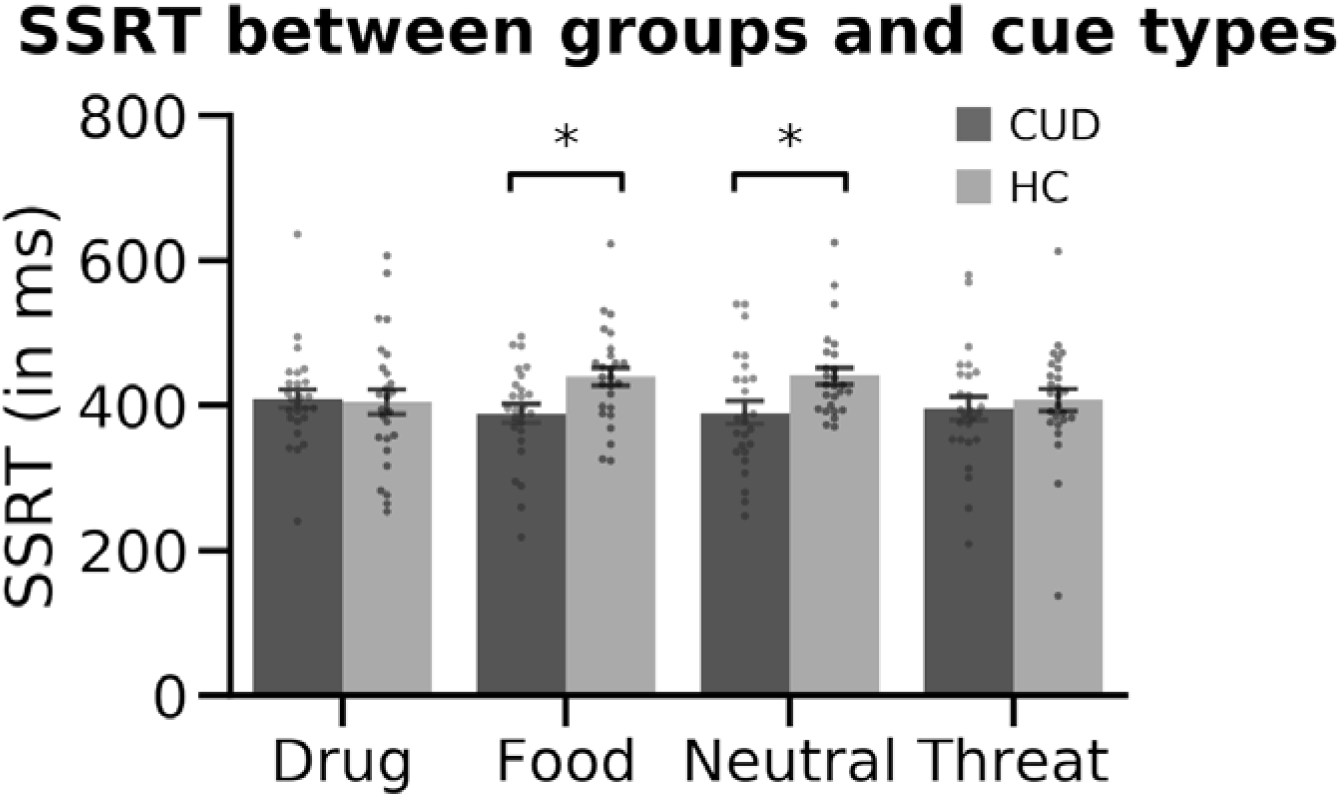
CUD and HC groups’ SSRT in response to each cue type. We found no significant main effects of Group or Cue but found a significant Group x Cue interaction (*p* = .027), where HC were significantly slower to stop than the CUD group in response to Food and Neutral cues. No drug-cue-related group differences were evident in SSRT.

The total score on the Severity of Dependence Scale positively correlated with overall SSRT (Spearman’s *r* = .40, *p* = .043, Holm-corrected), such that higher severity of cocaine dependence was associated with slower SSRT (worse performance). No other clinical measure showed a significant correlation with behavior (all *p*s > .05).

Due to missing data, we were able to extract post-task word rating data from 45 out of the 52 participants included in the analyses (CUD: n = 21; HC: n = 24). A two-way ANOVA with valence as the dependent variable, Group as between-, and Cue as within-subject factors revealed no main effect of Group, *F*(1,37) = 0.92, *p* = .343, a significant main effect of Cue, *F*(3,37) = 19.75, *p* < .001, and no Group x Cue interaction, *F*(3,37) = 0.25, *p* = .859. Pairwise comparisons of valence ratings between cue types across all participants indicated the following (from most to least positive): Food > Neutral > Drug > Threat, *t*(44) ≥ 7.27, *p* <.001. An identical two-way ANOVA with arousal as the dependent variable revealed no main effect of Group, *F*(1,37) = 1.64, *p* = .209, a significant main effect of Cue, *F*(3,37) = 5.53, *p* = .003, and no Group x Cue interaction, *F*(3,37) = 2.28, *p* = .095. Pairwise comparisons of arousal ratings between cue types across all participants indicated the following (from most to least arousing): Threat > Drug > Neutral, *t*(44) ≥ 2.58, *p* ≤ .013; Food cues, greater than neutral cues, *t*(44) = 2.33, p = .024, did not significantly differ from Threat or Drug cues (both *p*s > .05). Thus, across all subjects, drug words were rated as more negative than food words; their level of arousal did not differ.

### BOLD-fMRI results

#### Inhibitory control-related neural signaling (across all participants)

The whole-brain analysis of overall inhibitory control across all participants (i.e., neural signaling during Stop_Success > Stop_Fail regardless of cue type and group) yielded significant clusters in classical inhibitory control-associated regions, such as right vmPFC (Brodmann’s Area, or BA10), left OFC (BA47), left IFG (BA45), right frontal pole/dmPFC (BA8; among several other regions listed in Table 3, top). Next, we used mean SSRT (regardless of cue type) as a covariate to detect cue-general inhibitory control and found significant clusters namely in the right frontal pole/anterior PFC (BA10), right paracingulate gyrus (BA9), bilateral middle frontal gyrus/dlPFC (BA8 in both hemispheres), and among several others (see Table 3, bottom).

**Table 3.**
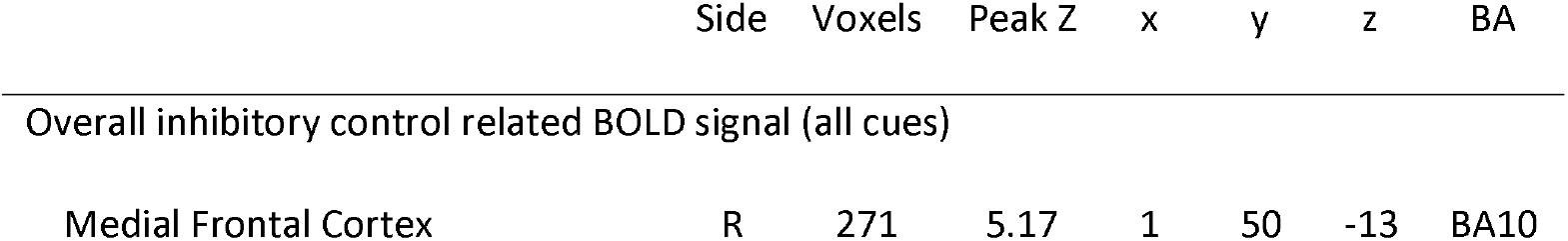
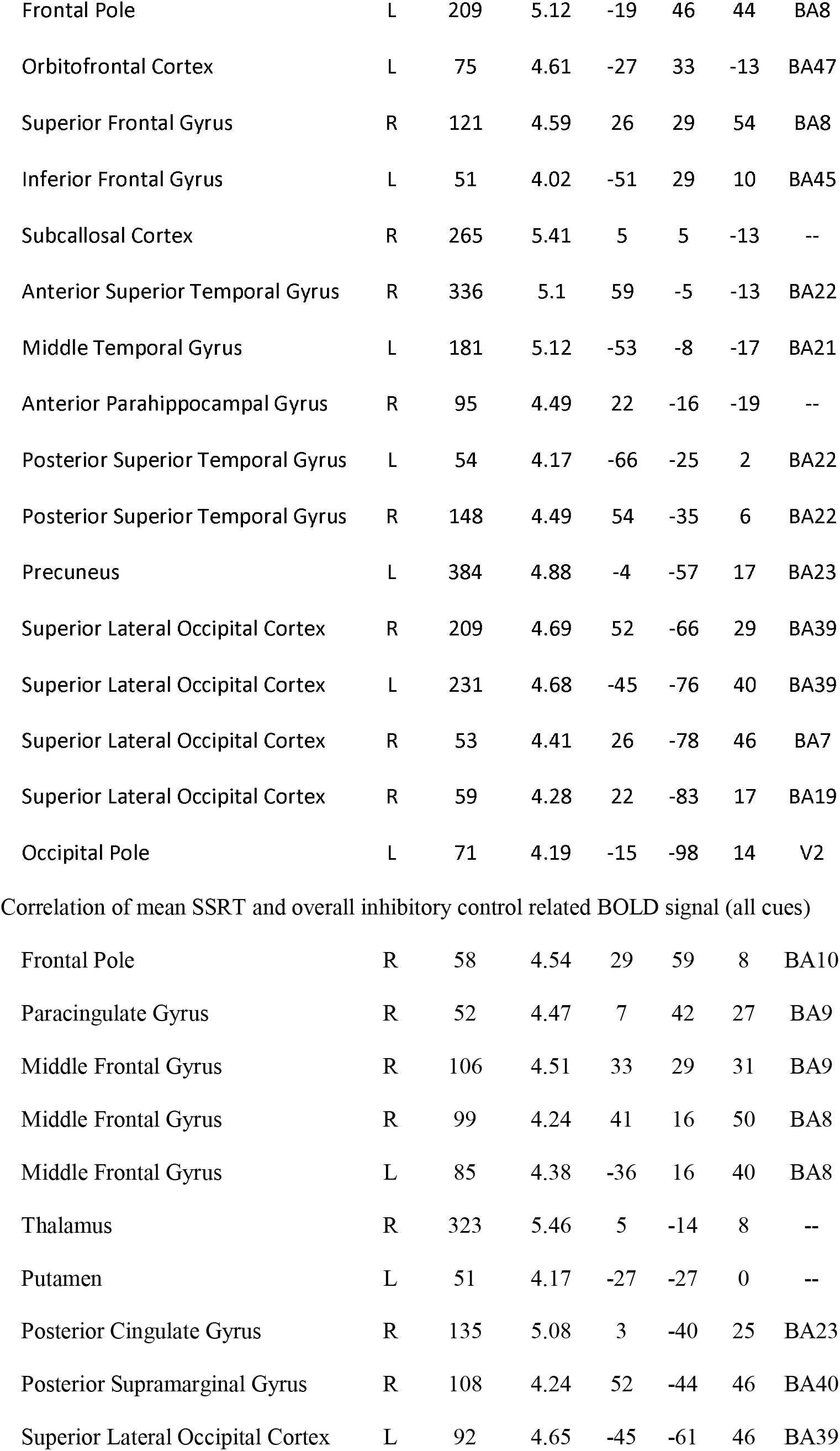
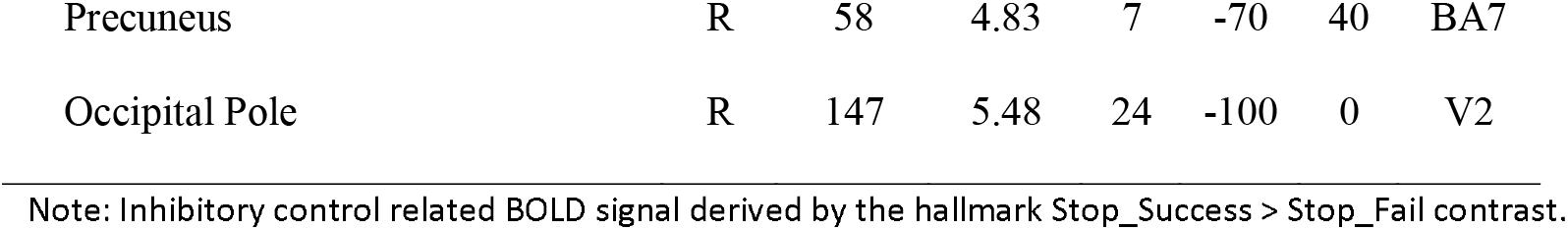
Inhibitory control related BOLD signal, all participants

|  | Side | Voxels | Peak Z | x | y | z | BA |
| --- | --- | --- | --- | --- | --- | --- | --- |
| Overall inhibitory control related BOLD signal (all cues) |  |  |  |  |  |  |  |
| Medial Frontal Cortex | R | 271 | 5.17 | 1 | 50 | -13 | BA10 |
| Frontal Pole | L | 209 | 5.12 | -19 | 46 | 44 | BA8 |
| Orbitofrontal Cortex | L | 75 | 4.61 | -27 | 33 | -13 | BA47 |
| Superior Frontal Gyrus | R | 121 | 4.59 | 26 | 29 | 54 | BA8 |
| Inferior Frontal Gyrus | L | 51 | 4.02 | -51 | 29 | 10 | BA45 |
| Subcallosal Cortex | R | 265 | 5.41 | 5 | 5 | -13 | -- |
| Anterior Superior Temporal Gyrus | R | 336 | 5.1 | 59 | -5 | -13 | BA22 |
| Middle Temporal Gyrus | L | 181 | 5.12 | -53 | -8 | -17 | BA21 |
| Anterior Parahippocampal Gyrus | R | 95 | 4.49 | 22 | -16 | -19 | -- |
| Posterior Superior Temporal Gyrus | L | 54 | 4.17 | -66 | -25 | 2 | BA22 |
| Posterior Superior Temporal Gyrus | R | 148 | 4.49 | 54 | -35 | 6 | BA22 |
| Precuneus | L | 384 | 4.88 | -4 | -57 | 17 | BA23 |
| Superior Lateral Occipital Cortex | R | 209 | 4.69 | 52 | -66 | 29 | BA39 |
| Superior Lateral Occipital Cortex | L | 231 | 4.68 | -45 | -76 | 40 | BA39 |
| Superior Lateral Occipital Cortex | R | 53 | 4.41 | 26 | -78 | 46 | BA7 |
| Superior Lateral Occipital Cortex | R | 59 | 4.28 | 22 | -83 | 17 | BA19 |
| Occipital Pole | L | 71 | 4.19 | -15 | -98 | 14 | V2 |
| Correlation of mean SSRT and overall inhibitory control related BOLD signal (all cues) |  |  |  |  |  |  |  |
| Frontal Pole | R | 58 | 4.54 | 29 | 59 | 8 | BA10 |
| Paracingulate Gyrus | R | 52 | 4.47 | 7 | 42 | 27 | BA9 |
| Middle Frontal Gyrus | R | 106 | 4.51 | 33 | 29 | 31 | BA9 |
| Middle Frontal Gyrus | R | 99 | 4.24 | 41 | 16 | 50 | BA8 |
| Middle Frontal Gyrus | L | 85 | 4.38 | -36 | 16 | 40 | BA8 |
| Thalamus | R | 323 | 5.46 | 5 | -14 | 8 | -- |
| Putamen | L | 51 | 4.17 | -27 | -27 | 0 | -- |
| Posterior Cingulate Gyrus | R | 135 | 5.08 | 3 | -40 | 25 | BA23 |
| Posterior Supramarginal Gyrus | R | 108 | 4.24 | 52 | -44 | 46 | BA40 |
| Superior Lateral Occipital Cortex | L | 92 | 4.65 | -45 | -61 | 46 | BA39 |
| Precuneus | R | 58 | 4.83 | 7 | -70 | 40 | BA7 |
| Occipital Pole | R | 147 | 5.48 | 24 | -100 | 0 | V2 |

Note: Inhibitory control related BOLD signal derived by the hallmark Stop\_Success > Stop\_Fail contrast.

#### Group differences in inhibitory control-related neural signaling

Whole-brain analysis of overall inhibitory control (regardless of cue type) between groups revealed decreased activation in the CUD group relative to HC in the left frontal pole/dmPFC (BA10: MNI space -4, 57, 21; peak Z = 5.04, 97 voxels) and left lateral occipital cortex (BA39: MNI space -42, -63, 27; peak Z = 4.16, 70 voxels; see Figure 3).

**Figure 3.**
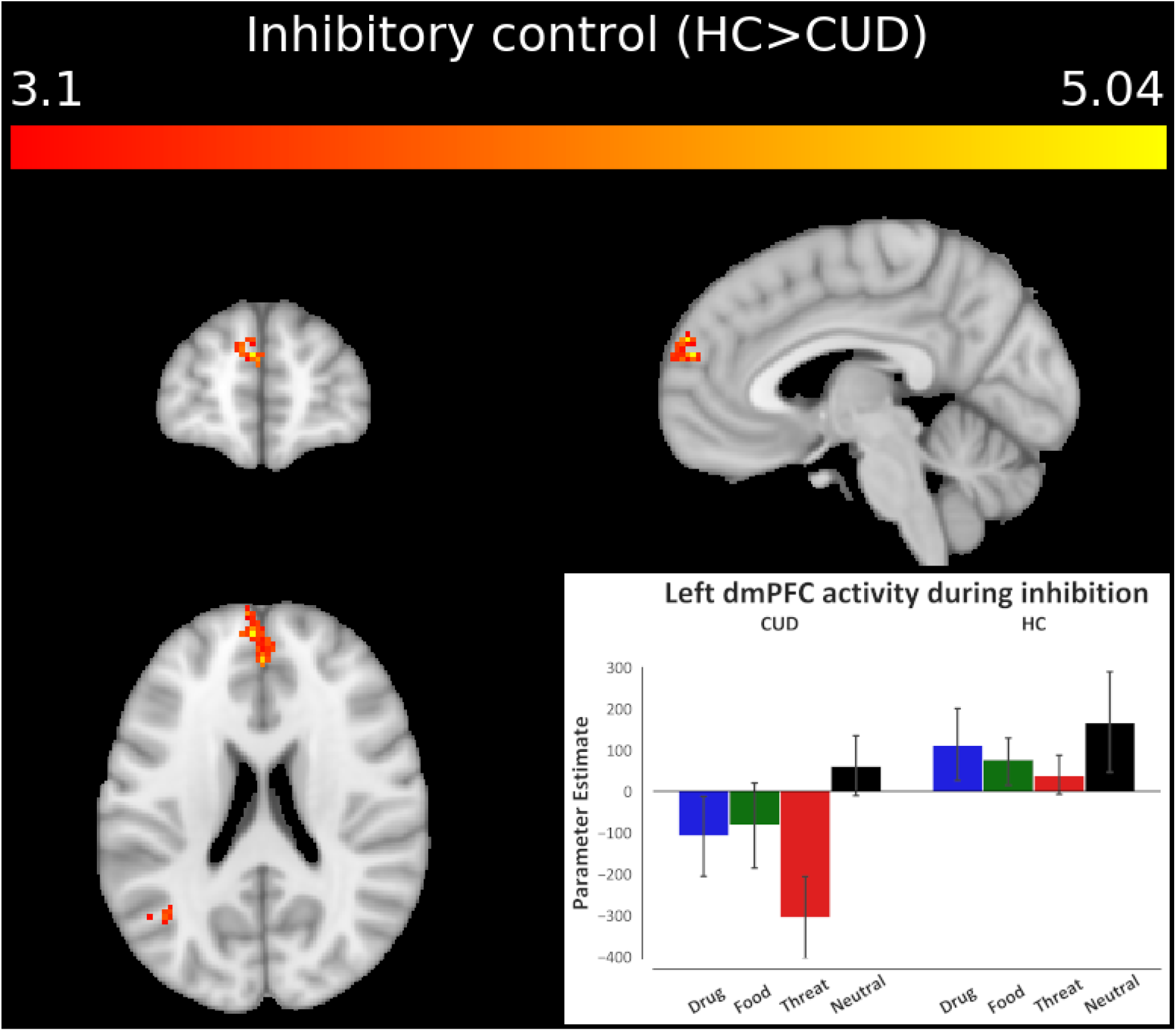
Brain activity differences across CUD and HC groups during inhibitory control, regardless of cue type. The analysis of overall inhibitory control revealed significantly lower activity in the left dmPFC in the CUD group compared to HC. Cluster defining threshold: Z > 3.1 (*p* < .001), cluster corrected at *p* < .05. The bar plot of activation patterns shows relative activation to all cue types within the voxels that display peak activation as a result of the overall inhibitory control contrast. Inhibitory control was modeled via the hallmark Stop_Success > Stop_Fail contrast of task events corresponding to all cues.

We also used mean SSRT as a covariate to determine potential group differences in the brain regions that correlated with stopping ability, and found significantly higher activations in CUD compared to HC in the posterior cingulate (BA31: MNI space -2, -35, 40; peak Z = 4.28, 54 voxels), left precuneus (BA7: MNI space -8, -66, 52; peak Z = 4.88, 105 voxels), and cuneal cortices (BA19: MNI space -23, -72, 19; peak Z = 4.22, 59 voxels), such that the higher the activity in these regions, the slower the SSRT (worse performance) specifically in CUD.

In sum, these results suggest that in CUD compared to HC the dmPFC is hypoactivated during inhibitory control, and greater activity in the posterior cingular/parietal regions is more closely associated with worse stopping ability.

#### Group differences in inhibitory control-related neural signaling under drug cue reactivity

Importantly, to capture the hypothesized iRISA interaction, we performed whole-brain analyses that focused on group differences in BOLD signal during drug versus food related inhibitory control, which allowed us to examine specific neural responses to drug cue reactivity compared to another salient pleasant (positively valenced) cue that is associated with consummatory behavior. The results revealed significant left dlPFC hypoactivations in the CUD group compared to HC during Drug > Food-related inhibition (BA8: MNI space -53, 12, 38; peak Z = 3.89, 62 voxels; see Figure 4), suggesting that specifically in CUD, this region exhibited decreased signaling during the inhibitory control of drug words (which were rated as more negatively valenced compared to food words in both groups). Similar analyses with Drug > Neutral-related inhibition did not yield significant results.

**Figure 4.**
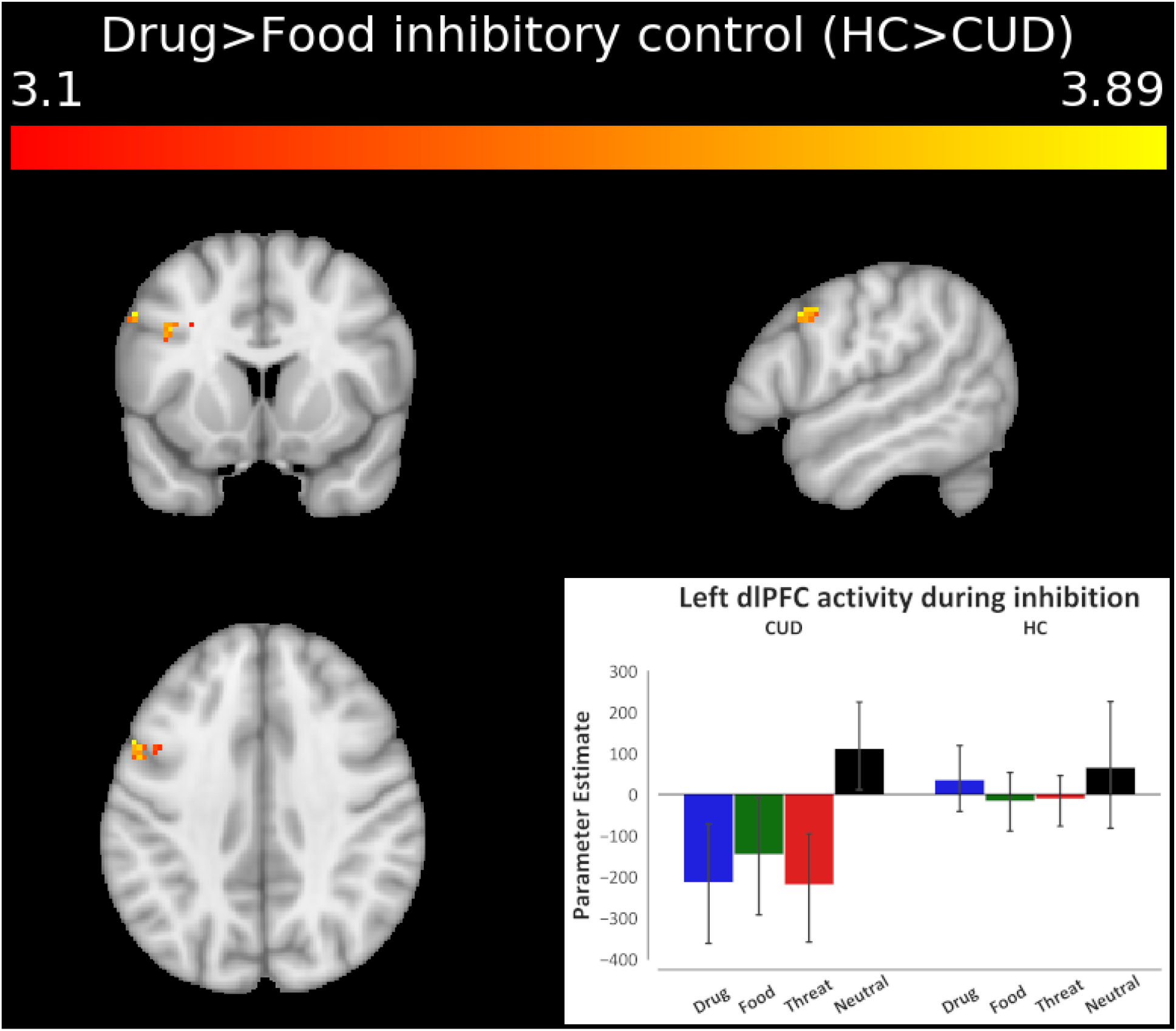
Brain activity differences across CUD and HC groups during inhibitory control under drug cue reactivity. The linear contrast of drug cue-versus a competing, non-drug positive reinforcer, food cue-related inhibitory control revealed significantly lower activity in the left dlPFC in the CUD group compared to HC. Cluster defining threshold: Z > 3.1 (*p* < .001), cluster corrected at *p* < .05. The bar plot of activation patterns shows relative activation to all cue types within the voxels that display peak activation as a result of the Drug > Food inhibitory control contrast. Inhibitory control was modeled via the hallmark Stop_Success > Stop_Fail contrast of task events corresponding to drug and food cues.

We also used Drug SSRT > Food SSRT as a covariate to determine group differences in which brain cue-sensitive inhibitory control regions correlated with Drug > Food-related stopping ability. A whole brain correlation found that the right superior lateral occipital cortex activity (BA39: MNI space 46, -66, 21; peak Z = 4.09, 61 voxels) negatively correlated with Drug > Food SSRT in HC compared to CUD, such that increased activity was associated with quicker stopping to drug compared to food cues in HC, and this correlation was significantly stronger than in the CUD group. Similar analyses with Drug > Neutral SSRT did not yield significant group differences in correlations.

#### Cocaine use severity and inhibitory control-related signaling under drug cue reactivity in the CUD group

CUD-focused analyses of drug use severity (the six cocaine use variables in Table 1) were conducted via unbiased, whole-brain correlations, effectively localizing the regions in which each of these drug use measures tracked the BOLD signal during drug-related (Drug > Food and Drug > Neutral) inhibition (see Table 4). Most notably, we found a significant positive correlation between recent frequency of use (days in the past month) and Drug > Food inhibition-related right OFC (BA47), right superior frontal gyrus/dmPFC (BA6) and left dlPFC (BA6) activity in the CUD group (see Figure 5). Using this same contrast (i.e., Drug > Food), a positive correlation was also observed with cocaine craving ratings in the right dlPFC (BA9), right IFG (BA44), and right middle temporal gyrus (BA21). Consistent with this direction of associations, negative correlations were observed with length of abstinence in the right anterior PFC (BA10), right dlPFC (BA44), and right precentral gyrus (BA6). Together, these results suggest that the more frequent the recent cocaine use, the higher the craving, and the shorter the abstinence, the higher the activity mainly in PFC regions during drug compared to food-related inhibitory control. Similarly, Drug > Neutral inhibition related BOLD signal positively correlated with recent cocaine use frequency in the left dlPFC (BA8) and cocaine craving in the left anterior PFC (BA8), left dlPFC (BA10), right dmPFC (BA6), left IFG (BA44), and the right hippocampus. Thus, similarly to the Drug > Food contrast, the Drug > Neutral contrast showed that higher prefrontal cortical activations (during drug compared to neutral-related inhibitory control) are related to more frequent cocaine use and higher craving.

**Table 4.** Whole-brain Drug > Food and Drug > Neutral inhibitory control related BOLD signal correlations with drug use measures, CUD group only

|  | Side | Voxels | Peak Z | x | y | z | BA |
| --- | --- | --- | --- | --- | --- | --- | --- |
| Drug > Food inhibitory control related BOLD signal |  |  |  |  |  |  |  |
| Past 30-day cocaine use |  |  |  |  |  |  |  |
| Orbitofrontal Cortex | R | 99 | 4.39 | 33 | 35 | -9 | BA47 |
| Superior Frontal Gyrus | R | 69 | 4.15 | 20 | 25 | 54 | BA6 |
| Middle Frontal Gyrus | L | 133 | 4.27 | -40 | 5 | 50 | BA6 |
| Cocaine craving |  |  |  |  |  |  |  |
| Middle Frontal Gyrus | R | 61 | 4.32 | 46 | 44 | 25 | BA9 |
| Inferior Frontal Gyrus | R | 71 | 4.2 | 44 | 14 | 25 | BA44 |
| Middle Temporal Gyrus | R | 153 | 4.54 | 52 | -5 | -21 | BA21 |
| Length of abstinence |  |  |  |  |  |  |  |
| Frontal Pole | R | 59 | -4.25 | 29 | 53 | 12 | BA10 |
| Middle Frontal Gyrus | R | 109 | -4.17 | 48 | 14 | 35 | BA44 |
| Precentral Gyrus | R | 163 | -4.42 | 35 | -5 | 67 | BA6 |
| Drug > Neutral inhibitory control related BOLD signal |  |  |  |  |  |  |  |
| Past 30-day cocaine use |  |  |  |  |  |  |  |
| Middle Frontal Gyrus | L | 66 | 4.16 | -42 | 14 | 46 | BA8 |
| Cocaine craving |  |  |  |  |  |  |  |
| Frontal Pole | L | 53 | 4.17 | -19 | 48 | 48 | BA8 |
| Middle Frontal Gyrus | L | 68 | 5.11 | -42 | 38 | 27 | BA10 |
| Superior Frontal Gyrus | R | 90 | 4.35 | 3 | 23 | 63 | BA6 |
| Inferior Frontal Gyrus | L | 88 | 4.82 | -53 | 23 | 25 | BA44 |
| Hippocampus | R | 55 | 4.21 | 22 | -20 | -13 | -- |
| Length of abstinence |  |  |  |  |  |  |  |
| No significant clusters |  |  |  |  |  |  |  |

Note: Inhibitory control related BOLD signal derived by the hallmark Stop\_Success > Stop\_Fail contrast.

**Figure 5.**
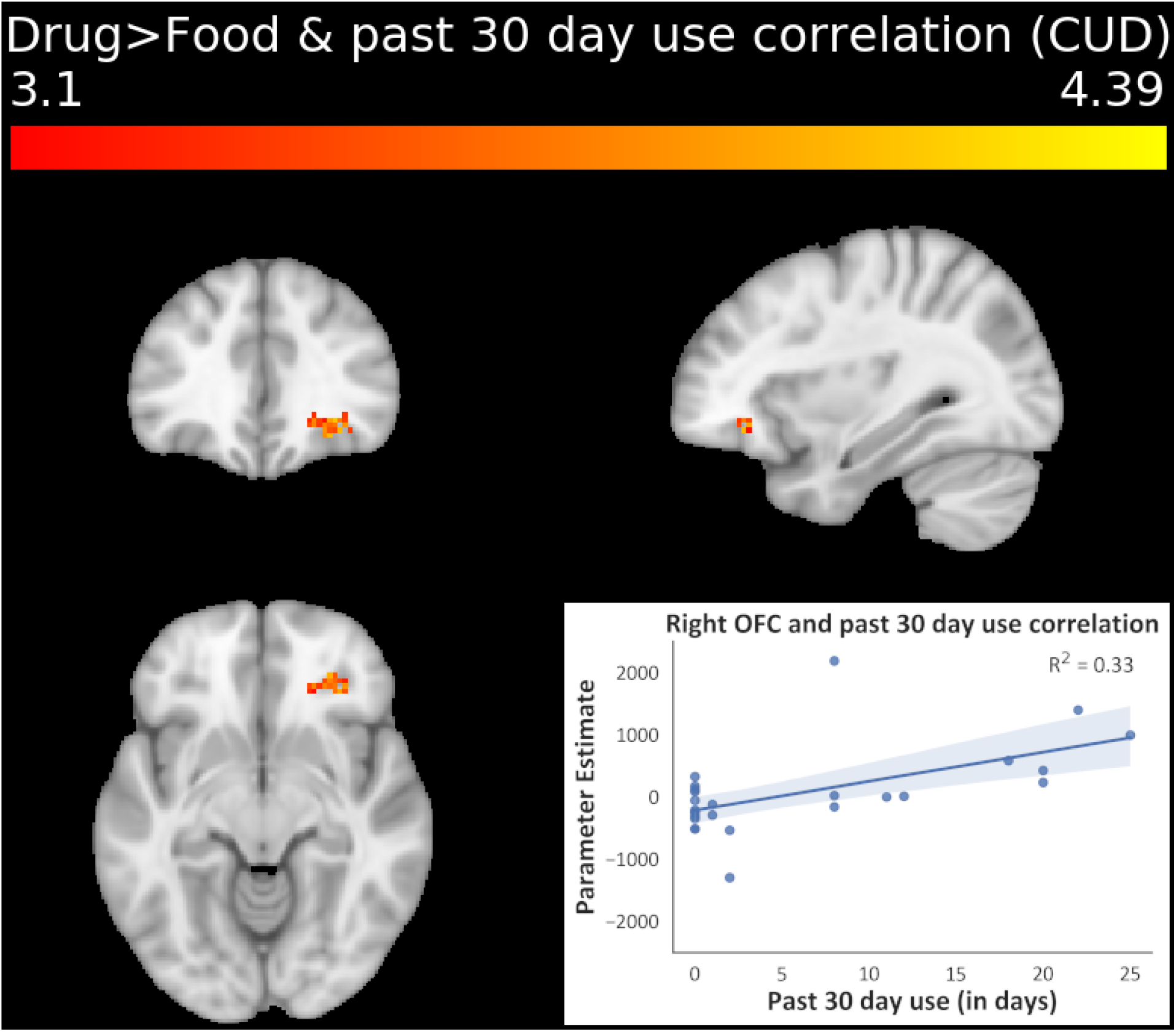
Whole-brain correlational analysis of cocaine use severity as assayed by recent cocaine use frequency in the past month, and its relationship with inhibitory control under drug cue reactivity (Drug > Food inhibitory control contrast). When entered into the GLM as a covariate in a whole-brain analysis, recent cocaine use frequency showed significant positive correlations with activity in the right OFC, right dmPFC, and left dlPFC in the CUD group, such that the more severe the cocaine use, the higher the activation in these prefrontal regions during inhibitory control under drug cue reactivity. Cluster defining threshold: Z > 3.1 (*p* < .001), cluster corrected at *p* < .05. Inhibitory control was modeled via the hallmark Stop_Success > Stop_Fail contrast of task events corresponding to drug and food cues.

## Discussion

Despite consistent evidence for inhibitory control and salience attribution impairments in individuals with substance use disorders [see reviews: (Zilverstand et al. 2018; Ceceli et al. 2021)], our understanding of how these hallmark addiction symptoms interact, and the neural underpinnings of this interaction, remains incomplete. Here, a novel fMRI SST permitted the neurobehavioral examination of the interaction between inhibitory control and salient drug and non-drug cue processing in CUD. Significant activations in the vmPFC, OFC, IFG, and dmPFC during successful compared to failed stops, independent of cue type or group, support our first aim to validate this novel task, showing its consistency with the PFC’s role in driving inhibitory control (Verbruggen and Logan 2008). Hypoactivations in the dmPFC during overall inhibitory control in the pre-selected group of individuals with CUD support our second hypothesis, contributing to the reliably documented body of work implicating decreased PFC signaling during a wide-range of neuropsychological functions, including response inhibition, in this population (Zilverstand et al. 2018). Third, and most importantly, we revealed evidence for the iRISA interaction in the CUD brain. Despite group similarities in both drug cue behavioral performance and arousal/valence ratings, and despite meeting the stringent criteria for SST’s horse-race model assumptions, compared to the HC group, CUD participants showed decreased dlPFC function during drug vs. food (a consummatory non-drug positive reinforcer) cue inhibition. The lower these dlPFC responses to drug vs. food cues, the lower the drug use severity encompassing less frequency of recent use, lower craving and longer abstinence.

The OFC and IFG activations during successful stopping in all participants regardless of cue type suggest that our novel task effectively captures inhibitory control in the brain. These regions are known to comprise the brain’s cognitive control network, with specific involvement in the suppression of inappropriate behaviors, especially during the SST (Aron and Poldrack 2006; Aron et al. 2007; Brockett and Roesch 2021). The significant dmPFC hypoactivations in CUD during overall inhibitory control agree with previous evidence where a similar dmPFC region was also hypoactivated in CUD compared to HC during inhibitory control—interestingly during Go/No-Go performance with neutral letter stimuli (Hester and Garavan 2004). Notably, the lack of behavioral performance deficits in CUD and comparable valence and arousal ratings between groups suggest that these PFC hypoactivations during inhibitory control are not driven by compromises in task comprehension, performance, or perception of stimuli. In general, while CUD may be expected to perform worse than HC in this task, evidence for (Fillmore and Rush 2002; Li, Milivojevic, et al. 2006; Wang et al. 2018) and against (Li et al. 2008; Vonmoos et al. 2013; Chao et al. 2019) prolonged SSRT in CUD vs. HC exists.

Agnostic to group and cue type, mean SSRT was positively correlated with dlPFC and paracingulate activity (among other regions), suggesting increased activation in these regions with slower stopping. This result disagrees with the direction of the correlation reported in several studies (Aron and Poldrack, 2006; Li et al., 2006a; Zhao et al., 2019; Chevrier and Schachar, 2020; Lee and Hsieh, 2017; but see de Wit et al., 2012). These studies largely reported a negative correlation between SSRT and cortical and subcortical activity (namely in the PFC and basal ganglia) including the default mode network—a set of brain regions that exhibit low-frequency oscillations at “rest”, typically associated with self-referential thought (Raichle et al. 2001). The source of the different direction of correlations is not clear but could be attributed to the variability introduced by the word stimuli, which may more readily elicit self-referential brain activity, especially in CUD. Indeed, compared to HC, in CUD the precuneus and posterior cingulate—two of the functional hubs of the default mode network (Raichle et al. 2001; Utevsky et al. 2014)—were more positively correlated with SSRT, suggesting that those who were slower to stop also exhibited higher default mode network activity during inhibitory control.

Most importantly, for the first time we demonstrate the neural signature of the iRISA interaction in CUD. In behaviorally comparable demographically-matched CUD and HC samples, we found evidence for decreased inhibitory control signal in the dlPFC only in CUD during drug cues relative to a non-drug reinforcer (food cues). One explanation for this dlPFC deactivation invokes the suppression of salient drug cue reactivity, in line with the dlPFC’s role in orchestrating craving, potentially by modulating the OFC, which combines the value of the craving-inducing cues with affective states and drug availability (George and Koob 2013), overall leading to decreased craving when the dlPFC is inactivated. Indeed, smokers who underwent dlPFC inactivation via transcranial magnetic stimulation (1 Hz) displayed reduced craving when cigarettes were immediately available, and dlPFC inactivation decreased craving-related OFC activity during a smoking cue-reactivity task (Hayashi et al. 2013). Adopting this perspective, our results suggest that to resemble the HC behavioral performance, the CUD group needed to decrease dlPFC signaling during inhibitory control especially during the potentially craving enhancing drug (vs. other salient) cues. Accordingly, those CUD individuals with the most severe drug use patterns (i.e., more frequent recent use, higher craving, and shorter abstinence) were less likely to exhibit decreased dlPFC (and OFC) response in the face of drug (vs. food) cues. It remains to be tested whether this pattern is a biomarker of resilience, whereby the ability to exert control during drug cue reactivity in the lab predicts craving and drug use outside the lab. An alternative explanation invokes self-medication (and vulnerability)—whereby those with more frequent recent cocaine use may be normalizing (increasing dlPFC activity, similarly to the HC) underlying neurocognitive dysfunction, masking (with cocaine, a stimulant) the neuropsychological deficits associated with cocaine addiction (Woicik et al. 2009; Parvaz et al. 2012, 2015). This latter interpretation is more consistent with the expected direction of effect for the dlPFC, whereby reduced activation during inhibitory control is a common marker of deficit. Accordingly, lateral occipital activity was associated with quicker SSRT in HC (but not CUD) in the drug context, which, combined with the relationship between slower SSRT and higher default mode network activity in CUD, alludes to a mechanism by which these non-PFC regions may be compensating by aiding performance in HC but not CUD. Inhibitory control-related comparisons between recent cocaine users and abstainers (or as a function of abstinence longitudinally) would be needed to contrast these perspectives in future efforts.

Limitations of the present study include high rates of data exclusion due to the stringent SSRT estimation parameters. While data filtering to validate SSRT is integral in capturing inhibitory control in the lab, performance feedback between task runs to remind participants of the instructions (e.g., ensuring that the participant does not wait for the stop-signal) may minimize the loss of data and statistical power (Verbruggen et al. 2019). Furthermore, although behavioral investigations show inhibitory slowing to emotionally salient picture (Verbruggen and Houwer 2007; Kalanthroff et al. 2013; Ding et al. 2020) and word stimuli (Herbert and Sütterlin 2011) in the general population, we found prolonged SSRT in response to food and neutral cues in HC, with no evidence for slowing for the other arousing cue types or in CUD. Thus, our task, which used words did not elicit the expected behavioral effect (e.g., slowing driven by drug words in the CUD). Supplementing these results by hunger and food craving ratings, and other relevant measures (e.g., time since last meal), is warranted in similar future studies. Using visual cues (e.g., drug use images or video clips) may have more effectively induced sustained cue-reactivity than word stimuli; however, the word stimuli were preferred for our trial-by-trial design and for rendering verbal features such as length and frequency uniform between conditions. These results should also be replicated with more sex-balanced samples that can inform about potential sex differences in the neural mechanisms of inhibitory control. Lastly, although we statistically accounted for neural responses to food and threat cues, in-depth analyses of these behavioral and neural patterns await samples that better represent relevant comorbidities (e.g., eating disorders, intermittent explosive disorder).

To the best of our knowledge, this is the first report of the brain systems that regulate inhibitory control under drug cue reactivity in human cocaine addiction. To better represent the addiction experience in humans, where reminders of cocaine use can exert control over behavior, we developed a novel SST to reveal the neural mechanisms of the iRISA interaction. Our results ascribe an important role to the dlPFC in the control of behavior under drug cue salience. Specifically, although the task promotes increased PFC function in cognitive control-related regions, drug vs. food cue-related hypoactivations were evident in CUD compared to HC, as associated with less frequent recent use, craving and with longer abstinence. This dlPFC drug cue suppression may be a marker of resilience (dampening craving-related processes) or a signature of vulnerability (where cocaine is used to normalize impaired cognitive processes in the short term, yet with long-term negative consequences). Together, our results point to the mechanisms behind lapses in self-control when faced with a salient drug context in addicted individuals. A closer examination with only treatment-seeking individuals is warranted to test whether these neurobiological patterns are related to deviations from treatment goals. Results could also offer the possibility to refine neuromodulation to mitigate the effects of craving on self-control and other drug-related behaviors in drug addiction.

## Data Availability

Available upon reasonable request as permitted by data sharing policies of the institutional review board.

## Acknowledgments

The authors declare no competing financial interests.

